# Comparison of a custom Photogrammetry for Anatomical CarE (PHACE) system with other Low- Cost Facial Scanning Devices

**DOI:** 10.1101/2023.04.16.23288631

**Authors:** Josiah K. To, Anderson N. Vu, Lilangi S. Ediriwickrema, Andrew W. Browne

**Affiliations:** Gavin Herbert Eye Institute, Department of Ophthalmology, University of California Irvine, Irvine California; Department of Biomedical Engineering, University of California Irvine, Irvine California; Institute for Clinical and Translational Science, University of California Irvine, Irvine California

## Abstract

**Purpose:** To compare a custom Photogrammetry for Anatomical CarE (PHACE) system with other cost-effective 3-dimensional (3D) facial scanning systems to objectively characterize morphology and volume of periorbital and adnexal anatomy.

**Methods:** The imaging systems evaluated include the low-cost custom PHACE system and commercial software product for the iPhone called Scandy Pro (iScandy) application (Scandy, USA), and the mid-priced Einscan Pro 2X (Shining3D Technologies, China) device and Array of Reconstructed Cameras 7 (ARC7) facial scanner (Bellus3D, USA). Imaging was performed on a manikin facemask and humans with various Fitzpatrick scores. Scanner attributes were assessed using mesh density, reproducibility, surface deviation, and emulation of 3D printed phantom lesions affixed above the superciliary arch (brow line).

**Results:** The Einscan served as a reference for lower cost imaging systems because it qualitatively and quantitatively renders facial morphology with the highest mesh density, reproducibility (0.13 ± 0.10 mm), and volume recapitulation (approximately 2% of 33.5 μL). Compared to the Einscan, the PHACE system (0.35 ± 0.03 mm, 0.33 ± 0.16 mm) demonstrated non-inferior mean accuracy and reproducibility root mean square (RMS) compared to the iScandy (0.42 ± 0.13 mm, 0.58 ± 0.09 mm), and significantly more expensive ARC7 (0.42 ± 0.03 mm, 0.26 ± 0.09 mm). Similarly, the PHACE system showed non-inferior volumetric modeling when rendering a 124 μL phantom lesion compared to the iScandy and more costly ARC7 (mean percent difference from the Einscan: 4.68 ± 3.73%, 9.09 ± 0.94%, and 21.99 ± 17.91% respectively).

**Conclusions:** The affordable PHACE system accurately measures periorbital soft tissue as well as other established mid-cost facial scanning systems. Additionally, the portability, affordability, and adaptability of PHACE can facilitate widespread adoption of 3D facial anthropometric technology as an objective measurement tool in ophthalmology.

**Precis:** We demonstrate a custom system for facial photogrammetry (Photogrammetry for Anatomical CarE -PHACE) to produce 3D renderings of facial volume and morphology which compares with more expensive alternative 3D scanning technologies.

## INTRODUCTION

Three dimensional (3D) digital surface reconstruction technologies and analyses have been increasingly applied to the fields of architecture, surveying, agriculture, sports, and medicine. In ophthalmology, two-dimensional photogrammetry has previously been used to evaluate and monitor tumor pathology, as well as follow a patient during the peri-operative period.^1–5^ The advantage of 3D facial anthropometric measurements is the ability to non-invasively measure, reconstruct, and analyze diverse pathologies that could affect clinical decision making. For example, continued advancements in 3D facial anthropometry have assisted in craniofacial flap planning and creation for cleft lip and palate repair, as well as helped to accurately quantify facial skin wrinkles and scars before and after laser resurfacing.^6–8^

These technologies typically fall under two categories: photogrammetry and structured light sensing. Photogrammetry is the 3D spatial reconstruction of shapes and features derived from the mathematical analyses of photographs acquired from multiple vergence angles.^9^ Objects in photographs are computationally aligned in 3D digital space to create object feature coordinates that are subsequently used to generate 3D surfaces.^9^ Structured light technologies, in contrast, project a known pattern of light onto a subject, which are then captured by cameras from two different angles of known deviation. Each camera captures differences in the projected pattern as the light becomes distorted while conforming to an object’s surface. Differences in pattern distortion are stored in reference matrices that correlate the objects’ camera coordinates to sets of relative spatial coordinates and are ultimately used to render a 3D surface model of the object.^10^

Available facial photogrammetry technologies include the 3dMD Face System (3dMD, Atlanta, GA), Vectra H1 or XT (Canfield Scientific Inc., Fairfield, NJ), and Eva (Artec 3D, Luxembourg). 3dMD products use structured light sensors and are considered “active” scanners as these devices contain a light source for optimized data capture. However, portability is limited due to large, multi-camera systems. The Vectra H1 and the Eva use photogrammetry and are “passive” scanners since subjects are illuminated using ambient light. Though less expensive than the 3dMD suite, these passive scanners have been less accurate due to sequential imaging and total duration of data acquisition.^11–13^ The 3dMD Face System costs approximately $25,000 USD, the Vectra H1 $13,000 USD and the Eva $15,000 USD.^14^ Although 3D anthropometry can augment standard clinical care and further inform medical decision making, technologies are not widely implemented due to cost, lack of portability, and training required for complicated scanning systems. Thus, we sought to compare and evaluate alternative low-cost 3D facial modeling systems for widespread clinical use.

## METHODS

This study received Institutional Review Board approval from the University of California, Irvine and was conducted in accordance with the Declaration of Helsinki. Studies performed were HIPAA-compliant and all participants enrolled provided written informed consent.

### Study Design

We compared a customized low-cost automated photogrammetric facial scanning system, termed the Photogrammetry for Anatomical CarE (PHACE) system, with other low- and mid-priced 3D scanning technologies that employed structured light to acquire scan data. 3D scanners that cost less than $1000 were considered low-cost and scanners more than $1000 but below $10,000 were considered mid-priced. PHACE and the iPhone Scandy Pro smartphone application (version 1.9.10, Scandy, USA) are low-cost systems, whereas the Einscan Pro 2X (Shining 3D Technologies, China) and Array of Reconstructed Cameras 7 (ARC7) facial scanner (Bellus 3D, USA) are two mid-priced structured light sensing scanners. Four tests were performed to characterize the utility and limitations of each technique for clinical use. To evaluate precision we quantified 3D model mesh density and reproducibility using both human subject models and a mannikin face mask. To evaluate scan accuracy we considered the Einscan 3D to be the reference test and compared the 3D model surface deviation and volume emulation from the other mid-price scanner and the two low-cost scanners. The mannikin was scanned in triplicate, and human subjects were scanned once with each of the 4 scanning modalities. Healthy adults of various Fitzpatrick skin pigmentation scores were included (mean age 30 ± 5 years; three males; one European, one East Asian, and one African descent). The manikin facemask was used to assess 3D scanning accuracy and reproducibility since it eliminated facial movement as a source of error and incorporated facial contours. All faces were scanned in the same environment, location, and by a single trained researcher.

### Facial Scanning Systems

The PHACE system has been previously described by To et al.^15^ Briefly, the PHACE system used off-the-shelf motorized turntables to rotate two Google Pixel 3 smartphones (Android 11 operating system) 360 degrees. Photographs were imported into Metashape (Agisoft, St. Petersburg, Russia) and an in-house computer script was used to construct and store photogrammetric 3D renderings on a local computer. The constructed photogrammetric 3D renders were exported as a wavefront object (.OBJ) into the 3D point cloud open source software CloudCompare (CC) where models were scaled and analysed^16^.

The handheld Einscan captured data using structured white light and the company’s proprietary software (EXScan Pro v3.4.05). Resolution was set to maximum (0.2 mm) and the scanner held approximately 18 inches from the subject. The scanner was rotated around the subject’s face approximately 225 degrees horizontally (ear to ear) and 140 degrees vertically (forehead to chin).

The iPhone Scandy Pro application utilized the built-in infrared (IR) structured light *True Depth* Camera system found on iPhone X models and newer versions. Scans for this study were acquired with an iPhone 11 with 4GB of RAM (iOS 14). Within the Scandy Pro application, resolution was maximized (0.5mm) and held approximately 10 inches from the face while scanning across the face in a path similar to the Einscan.

The Bellus ARC7 facial scanner captured data using an array of seven IR structured light cameras and processed the model within its proprietary software (ARC Scan App v1.8.12). Image acquisition and processing occured on a dedicated Windows Surface Pro 6 laptop. During imaging, the software instructed subjects to rotate their head left and right from shoulder to shoulder approximately 120 degrees horizontally. Rendered 3D models were exported using maximum resolution and minimal mesh smoothing.

### 3D Printed Phantom Lesions

Phantom lesions consisted of four custom 3D printed hemispheres used to simulate variable sized ocular and adnexal volumetric pathology. Each hemispheric lesion was printed using black polylactic acid filament (Hatchbox, California, USA) on a Prusa i3MKS (Prusa Research, Prague, Czech Republic) 3D printer with a layer accuracy of 150 μm. Subjects’ faces were scanned without the phantom lesions to establish a baseline facial mesh model. Faces were scanned a second time with the phantoms affixed with double-sided tape approximately 1 cm above the superciliary arch.

### Measurements and Analysis

3D models from each scanning technique were imported as a stereolithography (.STL) file into CC, where models were aligned, cropped, registered, and analyzed. Data rendering and analyses was performed on a Gigabyte Z390 Aorus Ultra Gaming PC running Windows 10 with an Intel Core i9-9900k 8-core CPU @ 3.6GHz, 48 GB of RAM, and a Nvidia GeForce RTX 2080 graphics card.

To analyze the mesh density, 3D scans of the manikin face acquired with the PHACE system, iScandy, and ARC7 were aligned to one of the Einscan models. Facial scans were further cropped to the same dimensions (1 mm x 1 mm) at the midline of the glabella. The mesh face density was subsequently calculated.

The reproducibility of each scanning technique was evaluated by measuring the model deviation between triplicate scans acquired on the same device of the mannikin face. The triplicate whole facial models were aligned, cropped, registered to each other, and then the distance deviation between each scan was measured. A quantitative color-coded depth map was used to represent the deviation between each model for both the periorbita and the entire face. Analysis of all models was performed for the entire face and for cropped regions, which only included the periorbital tissues.

3D Model accuracy was calculated by analyzing the surface deviation of the manikin 3D models produced by each scanning modality to a reference model rendered by the Einscan. Prior studies investigated the Einscan’s clinical accuracy by comparing rendered facial measurements with measurements from a vernier calipers, which showed no statistical differences.^17^ 3D models from each scanning modality were aligned and registered to the Einscan reference model and cropped to have identical boundaries. The absolute maximum deviation, root mean square (RMS), and distance mean in addition to standard deviation were calculated. A quantitative color-coded depth map represented each scanning technique’s regional accuracy of the mannikin’s face.

To quantify the volumetric accuracy of the phantom lesions, facial models with and without 3D printed phantom lesions were exported to CC. Models were manually aligned, automatically registered to each other using the iterative closest point function, and then exported into Meshmixer. Facial models without phantom lesions were Boolean subtracted from facial models with phantom lesions. Hemisphere reference diameters were measured using a digital caliper. Volumes were calculated for large, medium, small, and mini phantoms with known dimensions of 19.75 ± 0.04 mm (2157 μL), 9.78 ± 0.05 mm (260 μL), 4.72 ± 0.05 (124 μL), and 2.80 ± 0.04 mm (33.5 μL), respectively. Each rendered hemisphere’s volume was measured using Meshmixer’s analysis stability tool and compared with reference dimensions.

### Statistical Analysis

A Kruskal-Wallis analysis with multiple comparisons and a one-way ANOVA with Tukey’s multiple comparison test (IBM SPSS Statistics 27 (IBM, Armonk)) were used to compare RMS, distance deviation means, maximum mean absolute deviations, and mean volume percent difference to evaluate model reproducibility and accuracy. *p-*values less than 0.05 were considered to be significant.

## RESULTS

The advantages and disadvantages of each 3D reconstruction technology is presented in Table 1.

**Table 1.** Cost and Benefits - Different 3D imaging technologies applied to facial scanning

|  | **EINSCAN 3D SCANNER** | **PHACE** | **iScandy** | **BELLUS ARC7** |
| --- | --- | --- | --- | --- |
| Scanning Type | Structured white light | Photographs | Structured infrared Light | Photos and structured infrared light |
| Manufacturer Precision and Accuracy (mm) | 0.2 – 0.3 | 1 | 1 | 0.4 |
| Render Time (min) | 7 - 20 | 25 – 40 | Immediately | 1.5 |
| Cost (\$USD) | 5000 | 500 | 700 | 8000 |
| Portability | Moderate | Moderate | Easy | Difficult |
| Safety | Uncomfortable bright light | Safe | Safe | Safe |
| Accessibility | Proprietary hardware and software | Uses any mobile phone | Only on Apple iPhone | Proprietary hardware and software |
| Customizable | Low | High | Low | Low |
| Imaging Surface | Any surface (not black) | Any non-reflective surface | Any surface | Face only |

We compared mesh density (Table 2) of each scanning technology by acquiring triplicate scans of a manikin mask using each imaging modality. The Einscan created the most consistent models (lowest standard deviation between triplicate measurements) with the highest mesh face density. The PHACE system produced the fewest mesh faces with 6.5 times fewer mesh faces than the Einscan. Both the iScandy and ARC7 produced models with meshes about half the density of the Einscan. A statistical significance was seen with fewer mesh faces produced by each modality - PHACE, iScandy, and ARC7 compared to Einscan.

**TABLE 2.** Scanning Precision and Model Reproducibility of Mannikin Face Mask

| Scanning Modality | Mean Mesh Density | Whole Face Scans |  | Periorbital Anatomy Only |  |
| --- | --- | --- | --- | --- | --- |
|  |  | Mean Absolute Deviation (mm) | Mean Maximum Absolute Deviation (mm) | Mean Absolute Deviation (mm) | Mean Maximum Absolute Deviation (mm) |
| Einscan | 90.33 ± 3.51* | 0.01 ± 0.00, NS | 0.23 ± 0.04* | 0.01 ± 0.00, NS | 0.20 ± 0.03* |
| PHACE System | 13.33 ± 1.15* | 0.01 ± 0.01, NS | 1.76 ± 0.41* | 0.07 ± 0.03* | 0.86 ± 0.22* |
| iScandy | 42.67 ± 4.93* | 0.04 ± 0.04, NS | 1.71 ± 0.70* | 0.02 ± 0.01, NS | 1.06 ± 0.23* |
| ARC7 | 37.33 ± 1.15* | 0.01 ± 0.01, NS | 1.13 ± 0.25* | 0.04 ± 0.02, NS | 1.11 ± 0.25* |

\* $p$ -value $\leq 0.05$ ; NS, not significant

The reproducibility of each scanning modality is shown in Figure 1 and Table 2. The Einscan renders models with the highest reproducibility (lowest RMS, mean absolute deviation, and mean maximum absolute deviation) and precision (lowest standard deviation). The Einscan demonstrated better reproducibility compared to PHACE (*P* = .007) and iScandy (*P* = .001) for an inanimate face when analyzing either the whole face or periorbita alone but not compared to the ARC7 (*P* = .475). The iScandy demonstrated the lowest reproducibility when scanning the whole manikin face (highest mean absolute deviation, standard deviation, and RMS). The PHACE system demonstrated lowest reliability when modelling the nose. No statistically significant differences in reproducibility between PHACE and all other imaging modalities was observed when rendering the whole face.

**FIG. 1.**
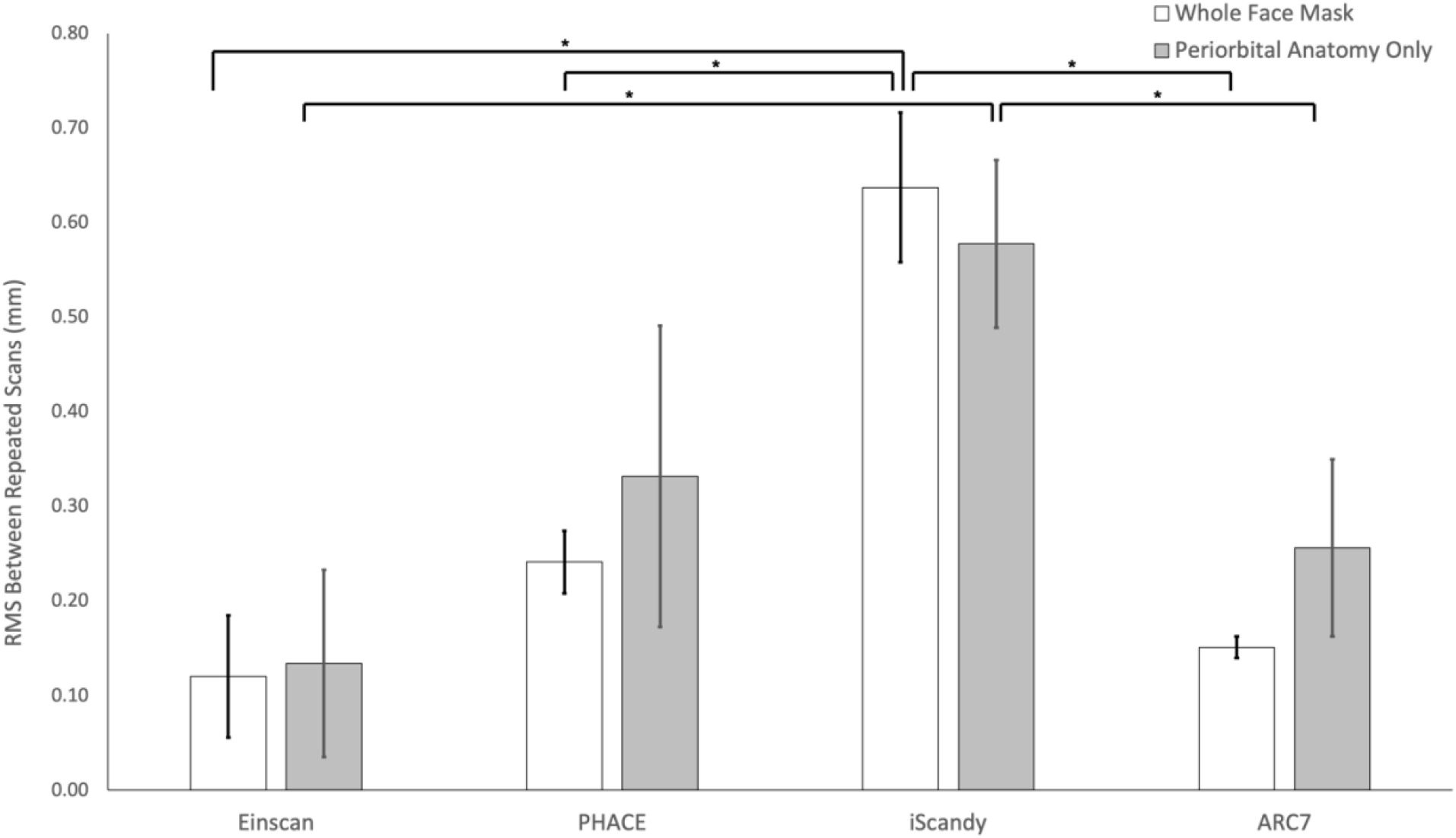
Comparison of the interscan RMS of three independently acquired scans of the same model to evaluate scanner reproducibility.

3D models of a manikin facemask were produced with each scanning technology (Fig. 2) where the accuracy and precision were subsequently compared using the Einscan model as a reference (Table 3). Displacement of 3D renders from the reference model are represented as quantitative deviation color maps (Fig. 2E-H). In the deviation maps, the color blue indicates inward deviation toward the center of the model, red indicates outward deviation from the model’s surface, and the color scale bar indicates the degree of deviation in millimeters. The color map produced for the Einscan model showed no deviation from itself (Fig. 2E) as it is the reference for the other modalities. The PHACE system modeled facial features similar to the iScandy except for the inward deviation along the nose (Fig. 2F) in the PHACE model and along the right parietal ridge (Fig. 2F) in the iScandy model. The iScandy and PHACE consistently created whole face 3D models with lower mean deviation and RMS (Table 3), which was also reflected by the deviation color maps (Fig. 2F – 2H). Models rendered from the ARC7 device often create facial models with significant outward deviation along the top of the head (Fig. 2H) and create false eyelid contours (2D white arrows) in the setting of absent globes in the manikin model (arrows in Fig. 2D). There were no statistically significant differences in periorbital and adnexal soft tissue mean deviation or RMS between all three modalities (Table 3) when compared with Einscan.

**TABLE 3.** Scanning accuracy using the Einscan model as a reference

| Scanning Modality | Whole Face Scans |  | Periorbital Anatomy Only Soft Tissue |  |
| --- | --- | --- | --- | --- |
|  | Mean Deviation (mm) | Mean RMS (mm) | Mean Deviation (mm) | Mean RMS (mm) |
| PHACE System | 0.01 ± 0.14* | 0.58 ± 0.10* | 0.03 ± 0.05, NS | 0.35 ± 0.03, NS |
| iScandy | -0.23 ± 0.15* | 0.69 ± 0.20* | 0.00 ± 0.01, NS | 0.42 ± 0.13, NS |
| ARC7 | 1.49 ± 0.50* | 3.04 ± 1.00* | 0.06 ± 0.03, NS | 0.42 ± 0.03, NS |

\* $p$ -value $\leq 0.05$ ; NS, not significant

**FIG. 2.**
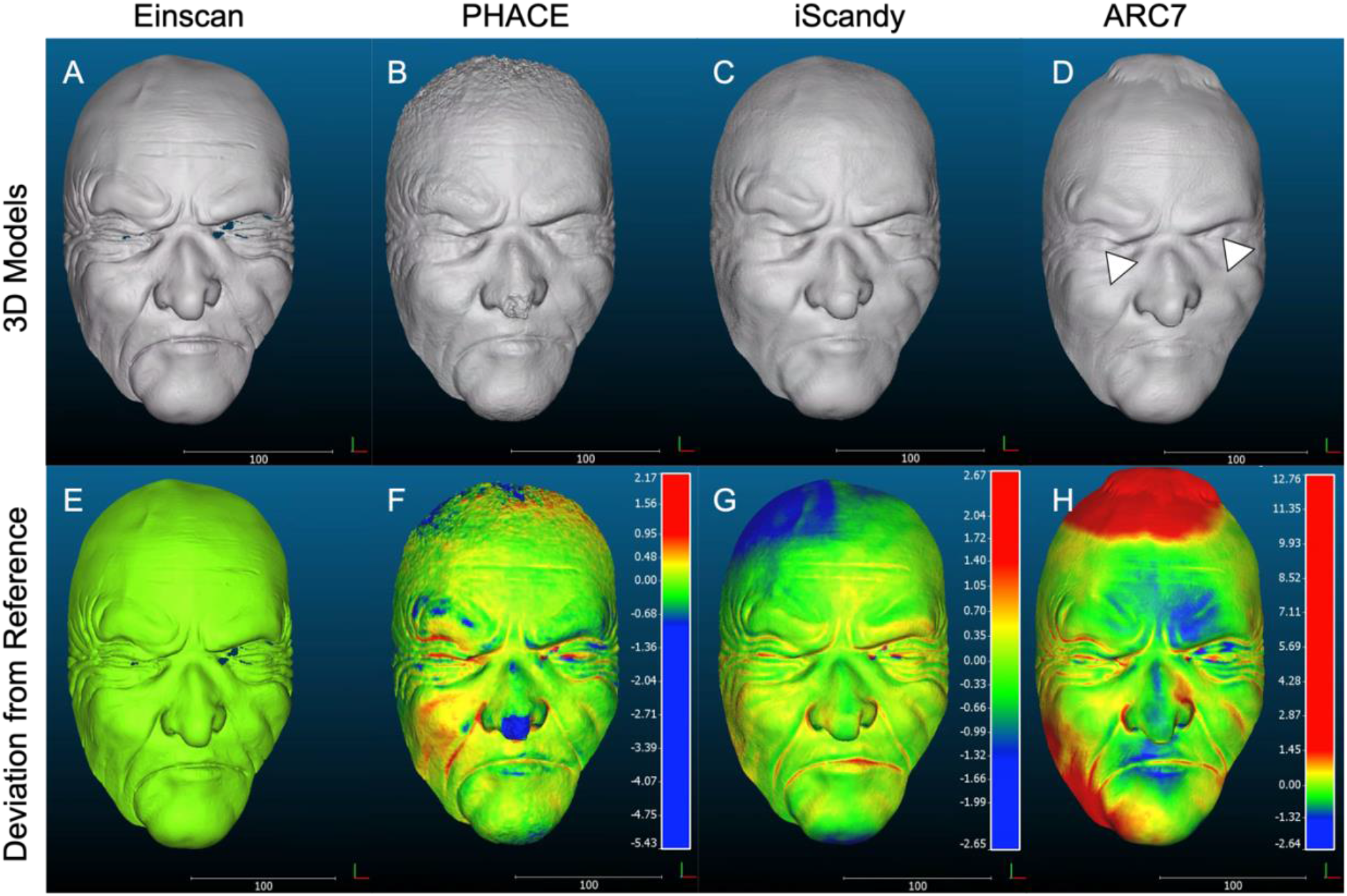
Comparison of 3D models of a manikin facemask produced with each scanning technology using the Einscan methodology as a reference. (A-D) Rendered mesh models of the manikin facemask. (E-H) Rendered models where color maps indicating amount of deviation from Einscan reference scan (A). Scale bar is in millimeters.

Digitally measured volumes of rendered dome-shaped phantom lesions affixed to the superciliary area (brow line) were compared with calculated volumes based on the diameters measured with a digital caliper. The Einscan qualitatively and quantitatively yielded models with the highest precision and accuracy (Fig. 3A and E). The quantitative comparison for volumetric measuremens of phantoms using Einscan, PHACE, iScandy, and ARC7 systems are shown in Figure 3E. There were no statistically significant differences between the PHACE, Einscan, and ARC7 systems for large, medium and small phantom lesions (Fig. 3E). The iScandy scanner rendered larger volumes for medium sized phantoms but all modalities overestimated the true volume of mini phantom lesions. The ARC7 produced models that partially recapitulated the mini phantoms (Fig. 3D).

**FIG. 3.**
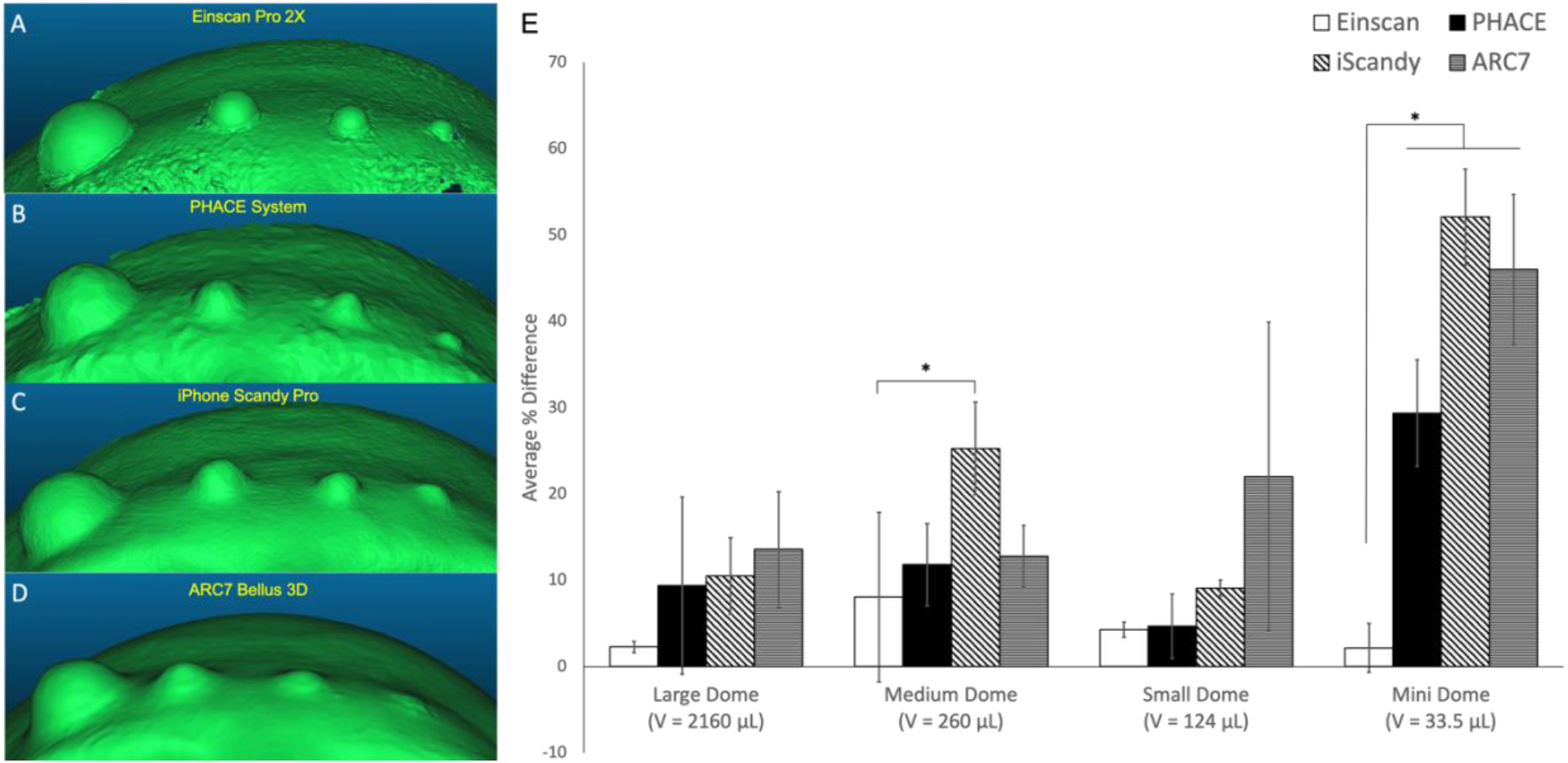
Worm’s eye-view of scanned and rendered 3D phantom lesions on participant 3 using the Einscan Pro 2X (A), PHACE system (B), iPhone Scandy Pro (C), and ARC7 Bellus 3D (D) of participant 2. (E), Comparison of each scanning technique’s average percent deviation when measuringwhite phantoms of various sizes placed along the superciliary arch on human subjects. *V* represents volume of each hemisphere.

The Einscan was unable to render models of human faces having different skin tones. The Einscan could only render the chin and portion of the cheeks of the subject with Fitzpatrick score 6 (Fig. 4A), as compared to the individual with Fitzpatrick score 2 (Fig. 4b). The model with Fitzpatrick score 6 (Fig. 4A) was missing facial data for for the ears, nose, orbit, periorbital adnexa, forehead and dark T-shirt worn by the subject.

**FIG. 4.**
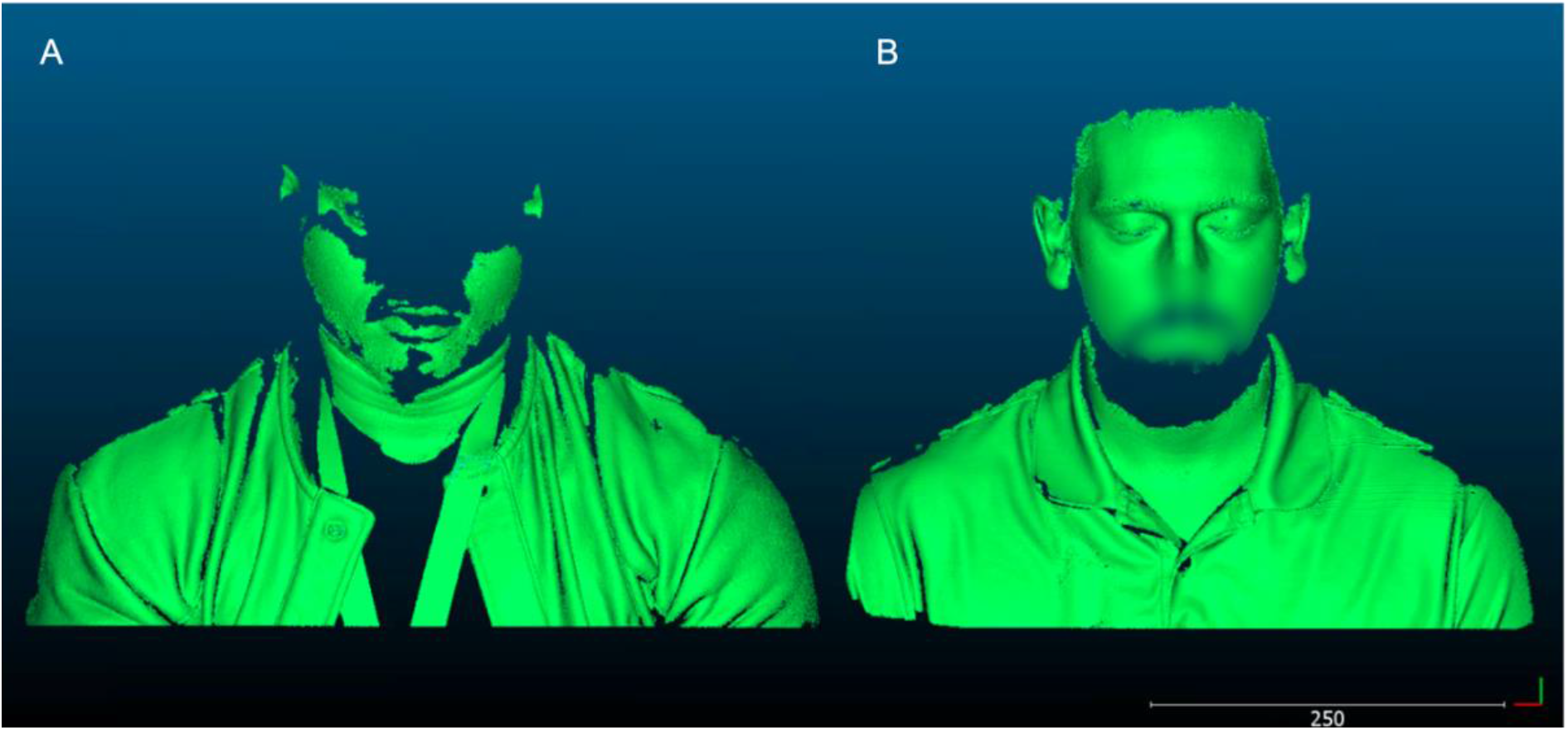
Comparison of 3D reconstructed models using the Einscan. Generated 3D model of a male with Fitzpatrick score 6 (A) and a male with Fitzpatrick socre 2 (B). Scale bar is in millimeters.

## DISCUSSION

3D anthropometry has been extensively applied to craniomaxillofacial surgery to evaluate surgical and clinical outcomes but its application to the periorbital region has been limited. ^18–29^ Recent technical advances have driven down the cost and access to powerful and portable 3D scanning systems.^14,30–33^ Systematic reviews have emphasized that although smart phone-based scanning systems remain less accurate, their variability remain within a clinically acceptable range for facial measurements as previously defined by an RMS value less than 2 mm from a standard reference.^31–34^

In this study, we experimentally compared low and mid-cost 3D scanning systems and discuss their clinical significance and applications. The Einscan Pro 2X was chosen to be the reference scanning device for accuracy analysis given prior validation of its clinical accuracy and precision.^17^ The accuracy of the PHACE system in our investigation performed better than a similar smartphone photogrammetry protocol presented by Nightingale et al.^30^ The iScandy results demonstrated less accuracy than Rudy et al’s iPhone X Scandy App study but still well within the clinically acceptable range (RMS < 2 mm).^14^ The limitation in Nightingale et al’s study was its exclusion of rendered data and analysis for the eyes and mouth, which, as our data from our human studies have demonstrated, are areas with the most variability due to small microexpressions. The reduced reproducibility of the PHACE system is likely related to perspective distortion from multiple camera vergence angles and magnitude of the depth difference between the nose and the remainder of the face. This may be overcome with telecentric lenses and greater distances between the subject and the camera. To the authors’ knowledge, the accuracy and efficacy of the ARC7 device has never been compared with other devices in a similar price range.

While published validation methods rely on displacement analysis and color depth maps to evaluate 3D model accuracy and precision of periorbital soft tissue, few studies used volumetry to investigate the accuracy of 3D anthropometric devices.^21^ Other than To et al, there are no studies to our knowledge that use volumetry to evaluate low-cost 3D facial scanning systems.^15^ Displacement analysis focuses on distances between mesh surfaces of a model and reference, which less clearly represent morphological changes seen more plainly with volume analysis. For example, volume analysis can more clearly depict quantiative changes in planar facial morphology that do not have depth changes such as a growing facial lesion. Therefore, it is important to not only measure the linear distance changes between analyzed and reference models but also measure the total *in vivo* volumetric changes. Our approach to assessing digital facial reconstruction techniques used phantom lesions made of 3D printed phantoms attached to the brow. Because the phantoms were printed with 100 μm resolution and measured with a digital caliper with 20 μm resolution, accurate volume calculations are possible since the error propagation due to measurement uncertainties remains lower than the resolution of each scanning modality. Accurate volume calculations permit the phantom lesions to be used as a known reference to compare measured volumes. Both the Bellus and iScandy had significant average percent differences from the calculated hemispheric volumes, which could possibly be due to a resolution limitation of structured IR light – as opposed to shorter wavelengths included in structured white light (Einscan) that discriminates facial details such as pigmented cutaneous features, rytids and larger contours. When a 3D model is generated, the mesh face density determines the smallest morphological features detectable by each scanning technique. The mesh face density has the potential to limit the model’s precision and accuracy. Differences in the number of projected structured light patterns from the Einscan, iScandy, and Bellus likely contributed to differences in model mesh density, with the Einscan projecting the highest density of structured light points. The PHACE system uses photogrammetric computation to infer mesh points from 80-120 images. The fewer number of data points compared to the density of structure light data may explain the low mesh density derived from PHACE models. The higher the mesh face density the lower the likelihood that it will be a limiting factor for model precision and accuracy.

The accuracy of each modality was assessed by analyzing the deviation between the rendered manikin face generated from the PHACE system, iScandy, and ARC7 compared to the Einscan reference model. Accuracy based on the RMS calculations indicated that the PHACE system and iScandy can statistically render a whole face model more accurately than the ARC7. However, color-coded depth maps more clearly indicated areas of inaccuracies, which gives users the opportunity to assess the utility and applicability of particular facial compartments. The ARC7 discrepancy in RMS between the whole face and periorbital tissue alone was due to the isolated but significant deviation along the superior coronal suture of the models. This deviation was possibly due to a combination of the IR light reflecting off the head of the manikin face as well as the need to turn the head during facial scanning acquisition, which may have led to software tracking inconsistencies. In isolating the periorbital soft tissue, both the RMS and color depth map indicated a high degree of accuracy and no statistically significant difference between the PHACE system, iScandy, and ARC7. Moreover, when adding both the quantitative and qualitative volumetry data to evaluate accuracy, the results indicated there are no statistically significant differences in accuracy between all three scanning modalities and each system could correctly render volumes as small as 124 μL. Therefore, the RMS, quantitative color-coded depth map, and volumetry data all indicated similarly clinically significant, accurate performance between the low-cost facial scanning systems and the ARC7.

The whole face quantitative color-coded depth map was useful to indicate inaccuracies of specific regions of facial models. The PHACE system inconsistently rendered prominent nose models, which was likely due to the limited image perspective data restricted due to the constrained angles that the cameras have with respect to the face. Moreover, in a prior characterization study of the PHACE system, the nose was inaccurately rendered in only two out of fifteen subjects.[To et. al] Therefore, in subjects with prominent noses cameras in the PHACE system could be positioned further apart to increase the perspective data for wider vergence angles.

### Clinical Applications

The Einscan 3D scanner was shown to most accurately and precisely recapitulate inanimate objects and human faces; however, it was notably limited to lighter skin tones, with Fitzpatrick scores 1-4. Because the Einscan Pro 2X used white structured light, an absence of reflected light from dark surfaces impeded data acquisition and model rendering. This limitation may be addressed with next generation systems produced by Shinging3D.

The PHACE system recapitulates facial morphology and volumetry as accurately as other low and mid-cost options and is independent of Fitzpatrick score. The PHACE system was the most cost-effective and accessible modality since it can use any smartphone. The total cost of the system is a few hundred dollars as opposed to a few thousand dollars for competing proprietary hardware and software. Additionally, minimal training was required as the data acquisition and model processing was nearly completely automated. The PHACE technique also includes the added benefit of producing a large photographic dataset from multiple angles for subsequent 2D image analysis if needed. However, the PHACE system required the most time to render a precise and accurate 3D model and occasionally modeled noses with prominent bridges inaccurately. In the clinic, the PHACE system is best suited for modeling facial morphology, depth, and volumetric changes of the orbit and adnexal tissues.

The iPhone Scandy Pro application was an affordable/cost-effective option that used high resolution IR structured light via the *True Depth Camera* found in newer generation iPhones (iPhone X and later). Furthermore, the iOS Scandy Pro app allowed all data acquisition, model processing, and storage to occur completely offline on the iPhone device without the need to send personally identifiable facial data to the cloud, unlike the Bellus3D iOS FaceApp. Additionally, the Scandy Pro app immediately generated models after scanning and consistently recapitulated most regions of the face, including the noses with prominent bridges. The iPhone Scandy Pro application is best suited for capturing whole face models in a situation that required a portable system such as at the slit lamp, bedside, and before/after surgery.

The ARC7 facial scanner acquired data and rendered a model in the shortest time between the four evaluated scanning modalities. Although the documentation was unclear as to how Bellus3D renders the final facial model, the company’s developer overview and API documentation indicate a facial tracking function using software.^35,36^ Therefore, it was possible that significantly atypical facial morphology that was not recognized by the software could reduce model accuracy. Moreover, the ARC7 is limited to imaging faces only. Therefore, the Bellus3D ARC7 facial scanner is suitable for medical contexts that require facial models to be created quickly, have grossly normal anatomical features, and only need anthropometric measurements from the forehead to the chin.

Overall, the PHACE system performed similarly to other low and mid-cost scanners and it is the most economical, accessible, and easy to use 3D anthropometric scanning tool. The iPhone Scandy Pro modeling technique is the most portable system that can consistently and quickly capture and render full face 3D models. The ARC7 is the fastest facial scanning system, whereas the Einscan is the most accurate and precise scanning system but exhibits limited use.

### Limitations

Limitations to this study can narrow the scope of clinical applicability. Subtle micro expressions drastically reduce the ability for standardized comparisons between models and ultimately lower the sensitivity of the analysis. In theory, after reconstructed models are registered to each other in 3D space, all changes detected are due to external factors altering the region of interest. In the clinical setting, changes between models will ideally be due to systemic or facial pathology, such as thyroid eye disease, trauma, burn, or infiltrative diseases that alter tissue volume and depth. However, facial micro-expression can alter the 3D model. Furthermore, when reconstructed models are registered in 3D space, subtle non-region of interest changes can alter the efficacy of the alignment and registration of the models to each other. Although the reconstructed models may be misaligned on the order of millimeters, the smaller the features being measured (e.g., a millimeter sized lesion) the greater the artificial change contributes to the error.

An additional limitation of this study was that all human subjects were imaged with their eyes closed because each of the scanning systems struggled to render transparent/translucent surfaces, such as the cornea. To obviate imaging the cornea directly, scans were taken with eyes closed so that the skin covering the orbit could be imaged to represent depth and volume of the cornea. Currently all light based photogrammetric and structured light systems also struggle with accurately capturing and rending periorbital tissue with eyes open. The next advances in 3D facial anthropometry development for periorbital soft tissue may benefit from methods to accurately represent transparent tissue such as the cornea.

In conclusion, we have qualitatively and quantitatively compared 3D facial models generated from affordable 3D reconstruction technologies. Implementing these techniques to study changes in the midface and orbital adnexa warrants further investigation to gain understanding of the value of these methods for objective facial anthropometry in a clinical setting.

## Data Availability

All data produced in the present study are available upon reasonable request to the authors

